# Is cognitive motor dissociation just a minimally conscious state “plus” by another name?

**DOI:** 10.1101/2025.06.11.25329346

**Authors:** Martin M. Monti

**Author notes:** Correspondence to: Martin M. Monti, Full address: UCLA Department of Psychology, Los Angeles, CA 90095, USA.

## Abstract

Cognitive Motor Dissociation (CMD) describes a condition whereby brain injury patients can demonstrate response to command through advanced electrophysiology and imaging assessments but are unable to do so through in standard, behavior-based, clinical assessments. Rightfully, significant emphasis has been placed on the fact that despite a similar behavioral phenotype, patients with CMD show better outcomes than patients without CMD. Yet, this finding is not overly surprising when considering that patients with CMD might *just* be minimally conscious state “plus” patients (MCS+; i.e., patients capable of response to command) who were misdiagnoses due to the known limitations of behavioral assessments in the presence of sensory, cognitive, or motor comorbidities.

The present work brings together 131 DOC patients, from two separate longitudinal studies, to assess whether patients able demonstrate response to command *via* brain responses but not behavioral responses (i.e., CMD patients) are “just” misdiagnosed MCS+ patients, in terms of short-term outcomes, or whether they represent a separate diagnostic entity. Robust general linear modelling reveals that, while CMD patients show greater short-term gains than patients with no evidence of CMD, consistent with prior work, these gains are not different from those seen in patients who can demonstrate response to command behaviorally (i.e., MCS+ patients). This pattern of results remains unchanged when separately analyzing Vegetative State (VS; i.e., entirely unresponsive) and Minimally Conscious State “minus” patients (MCS-; i.e., patients only able to show non-language-mediated response) with and without CMD, and when restricting analyses to traumatic brain injury patients only.

These findings suggest that, at least in terms of short-term outcomes, patients with CMD are not meaningfully different from MCS+ patients. Rather, CMD patients are best understood as MCS+ patients who were misdiagnosed likely due to the well-known limitations of behavioral assessments in the presence of comorbidities affecting sensory input, cognitive processing, and/or motor output. These results thus support the suggestion by the European Union practice guidelines to assign diagnoses based on the highest level of response obtained in a patient across behavioral and non-behavioral assessments, as well as the use of advanced assessments not only in behaviorally VS patients, consistent with the US guidelines, but also in MCS- patients. Finally, from a nosological perspective, these findings suggest that patients with CMD might best be described as “MCS+ patients with CMD,” to convey at once their true level of consciousness (i.e., MCS+) and the presence of motor output limitations (i.e., CMD).

## Introduction

Since the 2006 landmark case in which a behaviorally unresponsive brain injury patient, considered to be in a Vegetative State (VS), was shown to be able to voluntarily modulate their brain activity, as detected with functional magnetic resonance imaging (fMRI),^1^ the field of Disorders of Consciousness (DOC) has seen tremendous conceptual and technological progress, now codified in international guidelines.^2,3^ In large part, this work has been focused on developing novel means to detect ever more accurately the presence of consciousness, something that can be very complex in circumstances in which a patient might suffer from comorbidities affecting sensory input, cognitive processing, or motor output.^4^

In the pages of *Brain*, for example, Pan and colleagues,^5^ presented an electroencephalography (EEG)-based brain computer interface (BCI) protocol, capable of detecting the presence of voluntary attentional responses in patients unable to demonstrate response to command in conventional bedside clinical assessments such as the Coma recovery Scale Revised (CRS-R).^6^ In other words, Pan and colleagues have developed a novel approach to detecting the presence of Cognitive Motor Dissociation^7^ (CMD) – that is, a condition in which a brain injury patient is able to demonstrate response to command through EEG or fMRI based assistive technologies, but is unable to do so through motor output, likely due to a disconnection in thalamo-cortical circuits mediating voluntary motion.^8^ As remarked in the accompanying commentary,^9^ the contribution of Pan and colleagues has important diagnostic implications since it shows that 44% of patients who appeared unable to follow commands behaviourally could do so through a BCI system. Furthermore, the majority of these cases (53%) were patients with a Vegetative State diagnosis (sometimes referred to as Unresponsive Wakefulness Syndrome; UWS), thus considered to be unconscious, with the remainder being patients having a Minimally Conscious State “minus” (MCS-) diagnosis, implying they could only show subtle behavioral signs of awareness (e.g., eye-tracking), but no evidence of language comprehension or response to command.^10^ A particularly important and novel aspect of the work is that it addresses the question of whether conditions such as CMD are prognostically relevant. And indeed, Pan and collaborators show that VS and MCS- patients with CMD, at a 3-month follow-up, show greater likeliness of being in a higher level of consciousness (as measured by diagnostic category and CRS-R score, respectively) compared to their counterparts showing no evidence of CMD.

As exciting as this findings is, asking whether patients with a VS diagnosis found to have CMD are more likely to show signs of recovery compared to VS patients who do not show any evidence of CMD, despite their indistinguishable behavioural phenotype,^9^ is important, but incomplete. In what follows, I leverage the data presented in Pan *et al.* and integrate it with data published by Giacino, Whyte and collaborators,^11^ to addressing two additional important – yet controversial – questions. First, it seems intuitive that VS patients (as defined behaviorally) who demonstrate CMD would show greater ameliorations compared to VS patients not showing CMD. For, by virtue of showing response to command, if *via* brain activity, patients with CMD are revealing that they are MCS+ patients who were “mistaken” for VS patients due to the well-known limitations of behavioral assessment.^12–17^ Seen in this light, finding that patients who can perform response to command have better outcomes than VS patients is a well-established finding.^18^ The crucial question, then, is whether patients with CMD are meaningfully different in terms of prognosis from MCS+ patients (i.e., patients who can demonstrate response to command behaviorally). In other words, are patients with CMD “just” MCS+ patients by another name? Second, the present work addresses the question of whether advanced technologies such as that presented by Pan *et al.* should be applied in the routine management of DOC patients, a controversial^19^ issue that separates the US^2^ and EU^3^ guidelines from the UK^20^ ones, and whether they should be employed in both VS and MCS- patients,^21^ an issue upon which the US and EU guidelines fail to align. The answer to these questions has both very practical implications concerning the appropriate management for DOC patients as well as broader ramifications with respect to the appropriate nosology of disorders of consciousness.

## Materials and methods

### Data

Data associated with the experiment reported by Pan *et al*^5^ were collected from the Supplementary Table 1 associated with the paper. In particular, the total CRS-R scores at baseline and at the 3-month follow-up were extracted, together with the initial diagnosis associated with each patient (i.e., their behavioral phenotype), whether they showed a positive response to the BCI paradigm, as well as time since injury (TSI), age, sex, and etiology. Since the paper by Pan and colleagues did not include a comparison sample of MCS+ patients, their data were supplemented with longitudinal CRS-R scores of a subset of the patients presented by Giacino, Whyte and collaborators in the context of an interventional Amantadine trial.^11^ Specifically, for patients who appeared to be MCS+ at enrollment, CRS-R total score at baseline and at the last follow-up were extracted, together with each patient’s TSI at enrollment, age, sex, and etiology.

### Analysis

A total of 131 observations were extracted from the two papers (78 from Pan *et al*., and 53 from Giacino, Whyte, *et al.*; see Table 1). Baseline CRS-R total score, change in CRS-R total score at follow-up, age, sex, time since injury (TSI), initial diagnosis (i.e., VS, MCS-, MCS+), and, for the Pan *et al.* data only, CMD status were all included in the analysis. Since quantitative variables were not normally distributed, as assessed with a Shapiro-Wilk test of normality (W=0.614, p<0.001; W=0.943, p<0.001; W=0.923, p<0.001; W=0.931, p<0.001; for TSI, age, CRS-R total score at baseline, and CRS-R total score change, respectively), the following analyses use robust variants of the conventional general linear model methods, as described below. Analyses were performed in Jamovi 2.6.44^22^ (with the robust general linear model analyses implemented through GAMLj^23^).

**Table 1.** Demographics and clinical information for the included sample by group (no CMD, CMD, MCS+). (Abbreviations: Med, median; SD, standard deviation; Min, minimum; Max, maximum; CMD, cognitive motor dissociation; MCS+, minimally conscious state +; TSI, time since injury; CRS-R, Coma Recovery Scale Revised.)

|  | Group | N | Mean | Med | SD | Min | Max |
| --- | --- | --- | --- | --- | --- | --- | --- |
| **Age (years)** | **no CMD** | 44 | 37.82 | 37.00 | 15.149 | 15.000 | 70.00 |
|  | **CMD** | 34 | 37.94 | 41.50 | 13.303 | 15.000 | 62.00 |
|  | **MCS+** | 53 | 38.34 | 38.03 | 16.409 | 16.263 | 64.80 |
| **TSI (months)** | **no CMD** | 44 | 4.19 | 3.50 | 3.438 | 1.000 | 20.00 |
|  | **CMD** | 34 | 3.51 | 3.00 | 2.930 | 1.000 | 18.00 |
|  | **MCS+** | 53 | 1.58 | 1.33 | 0.673 | 0.900 | 3.57 |
| **CRS-R total (baseline)** | **no CMD** | 44 | 7.00 | 7.00 | 2.102 | 4 | 12 |
|  | **CMD** | 34 | 7.62 | 7.00 | 2.202 | 3 | 13 |
|  | **MCS+** | 53 | 13.17 | 13.00 | 3.683 | 6 | 20 |
| **CRSR total (follow-up)** | **no CMD** | 44 | 8.00 | 7.00 | 2.812 | 4 | 17 |
|  | **CMD** | 34 | 13.65 | 14.00 | 4.984 | 6 | 23 |
|  | **MCS+** | 53 | 19.30 | 22.00 | 5.301 | 2 | 23 |
| **CRS-R change** | **no CMD** | 44 | 1.00 | 0.00 | 2.057 | 0 | 10 |
|  | **CMD** | 34 | 6.03 | 4.50 | 4.455 | 0 | 20 |
|  | **MCS+** | 53 | 6.13 | 6.00 | 5.299 | -14 | 15 |

First, two robust ANOVAs were run to assess group differences in age and time since injury, together with a χ^2^ test of association to assess the distribution of male and female patients across the three groups.

The main analysis employed a general linear model to assess the association between group and change in CRS-R total score between baseline and follow-up while controlling for demographic and clinical variables. Specifically, the model included main effects of group (no CMD, CMD, MCS+), sex (male, female), time since injury (TSI; in months), age (in years), baseline CRS-R total score, and the group by sex interaction. Robust standard errors were calculated with the HC3 method,^24^ due to a significant Levene test for homogeneity of variances, while confidence intervals were estimated with bootstrap-based (BCa) method^25^ to account for non-normality in the variables’ distributions. Follow-up pairwise post-hoc comparisons to assess effects across groups were conducted on estimated marginal means (i.e., on means adjusted for all other included variables) and corrected for multiplicity using the Bonferroni-Holm stepdown procedure.^26^

To assess the central question at a greater level of granularity, the general linear model above was repeated separating VS and MCS- patients, thus resulting in a 5-level grouping variable (i.e., VS no CMD; VS with CMD; MCS- no CMD; MCS- no CMD; MCS+). All other aspects of the analysis remained unchanged.

Finally, because of an imbalance in aetiology between the two samples of patients, with VS and MCS- patients presenting a mix of traumatic (TBI; 52%), cardiovascular (CVD; 26%), and anoxic (AIB; 22%) brain injuries as compared to MCS+ patients, all presenting traumatic injuries, the main analysis was repeated, again, including only TBI patients. All other aspects of the analysis remained unchanged.

As determined by the UCLA IRB, the work described here is not research involving human subjects as defined by DHHS and FDA regulations.

### Data availability

The data from Pan *et al*.^5^ are publicly available online as a supplementary table in the original article: https://oup.silverchair-cdn.com/oup/backfile/Content_public/Journal/brain/143/4/10.1093_brain_awaa026/2/awaa026_supplementary_data.pdf The data from Giacino, Whyte, *et al.* were provided by the authors of that work by signed permission. Application for access to those data should be requested with the authors of that work.

## Results

The analysis included all 78 patients described in Pan *et al.*^5^ as well as 53 patients from the sample in Giacino, Whyte *et al*. ^11^ As shown in Table 1, this sample included 45 VS patients, of which 18 demonstrated CMD, 33 MCS- patients, of which 16 demonstrated CMD, and 53 MCS+ patients. As assessed by separate robust ANOVAs (using the median method), the three groups (no CMD, CMD, MCS+) did not differ on age (F(2,128)=0.015, p=0.985). Conversely, as expected, the three groups did differ in terms of time since injury (F(2,128)=14.0, p<0.001), with post-hoc pairwise tests indicating that the MCS+ group was assessed at significantly shorter time since injury compared to both the CMD group (difference = 1.94 months, p < 0.001, d = 0.77) and the no CMD group (difference = 2.62 months, p < 0.001, d = 1.04), whereas there was no significant difference between the CMD and no CMD groups (p = 0.355). Indeed, estimated marginal means were 4.19 months [95% CI: 3.31–5.35] for the no CMD group, 3.52 months [2.75–4.62] for the CMD group, and 1.58 months [1.39–1.77] for the MCS+ group. Finally, no significant association was detected between group and sex (χ²(2)=0.843, p=0.656).

The main general linear model analysis shows that group (i.e., no CMD, CMD, MCS+) was a significant predictor of CRS-R change at follow-up (F(2,122) = 17.705, p < 0.001, ω^2^_p_=0.224). In addition, baseline CRS-R total score was also found to be a significant predictor of change (F(1,122)=4.293, p=0.040, ω^2^_p_=0.039), with higher initial scores associated with smaller subsequent gains (β=-0.278, t(122)=-2.072, p=0.040). No significant effects of sex, age, TSI, or the interaction between group and sex were observed (all p > 0.05). Post-hoc comparison of the groups’ estimated marginal means (i.e., the means adjusted for time since injury, sex, age, baseline CRS-R total score) show that patients with CMD have greater CRS-R changes at follow-up compared to patients who did not show any evidence of CMD (t(122)=4.18, p*_holm_*<0.001, Cohen’s *d* for the model-adjusted means [d_mod_]=1.190), with the CMD group having an estimated CRS-R change marginal mean of 5.248 points (95% CI: 3.333–7.539) versus an estimated marginal mean of 0.255 (95% CI: - 0.877–1.178) for the no CMD group. Similarly, MCS+ patients exhibited an estimated marginal mean of 7.288 (95% CI: 5.190–9.000), also significantly greater than the no CMD group (t(122)=5.19, p*_holm_*<0.001, d_mod_=1.676). Crucially, however, the post-hoc pairwise comparison failed to uncover any significant difference between the CMD and the MCS+ groups (t(122)=1.31, p*_holm_*=0.192, d*_mod_*=0.486; see Figure 1a).

**Figure 1.**
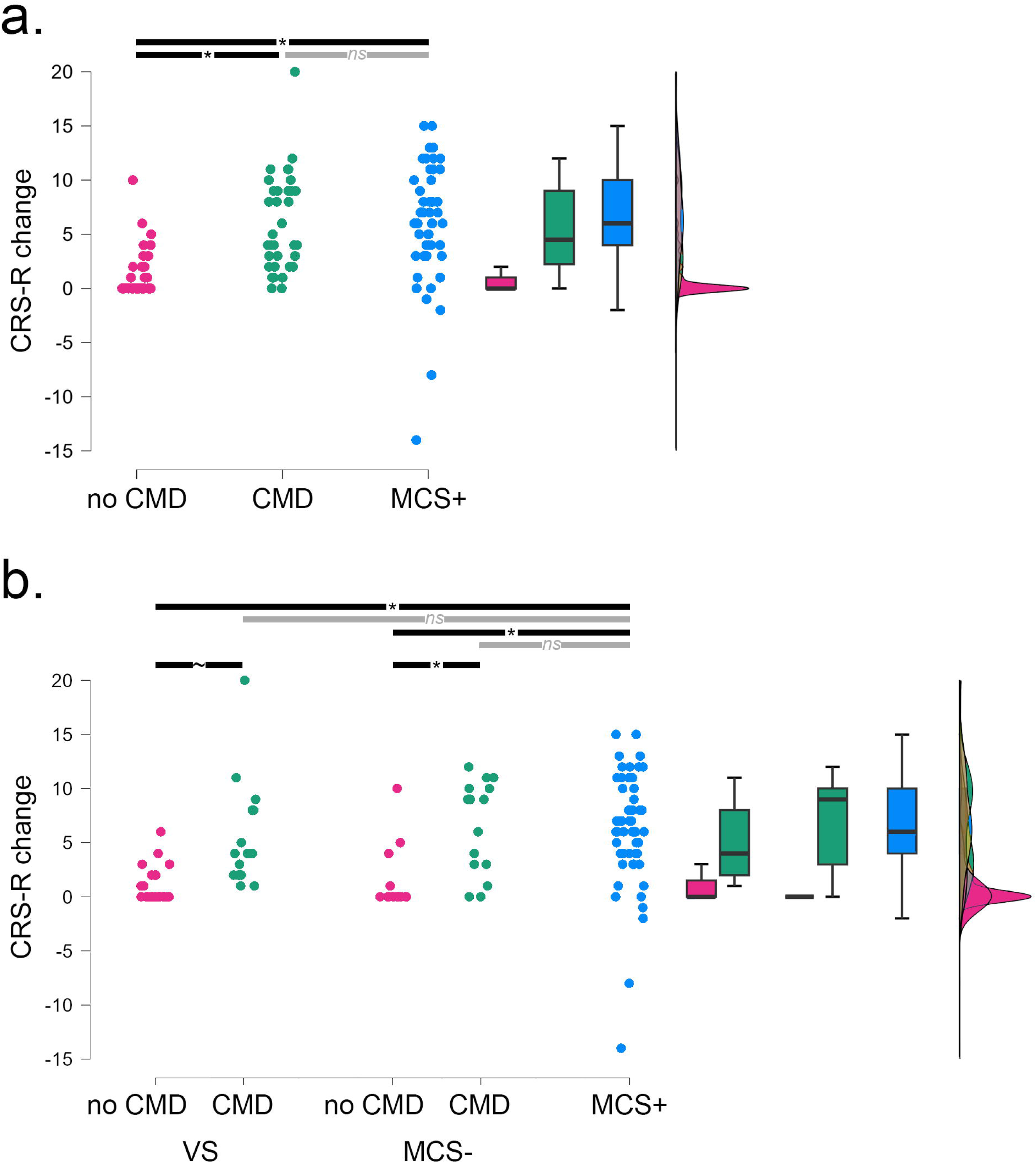
Raincloud plots for the change in CRS-R total score between baseline and follow-up. (**a**) Raincloud plot showing CRS-R total score change for the no CMD, CMD, and MCS+ groups. (**b**) Raincloud plot showing CRS-R total score change for the VS no CMD, VS with CMD, MCS- no CMD, MCS- with CMD, and MCS+ groups. In both panels, pairwise comparisons display the significance observed in the post-hoc analyses described in the text. (‘*’ indicates a significant difference in estimated means; ‘∼’ indicates trend towards significance; ‘ns’ indicates non-significant.)

To add further granularity to the comparisons, the above analysis was repeated after separating the VS and MCS- patients, thus resulting in a 5-level group variable (i.e., VS no CMD; VS with CMD; MCS- no CMD; MCS- with CMD; MCS+; see Table A1, in the Appendix, for a breakdown of demographics and clinical variables for each group). The general linear model again demonstrated a significant effect of group (F(4,118)=9.010, p< 0.001, ω^2^_p_=0.249) and baseline CRS-R (F(1,118)=5.874, p=0.017, ω^2^_p_=0.064), with no significant effect of time since injury, sex, age, or the interaction of sex and group (all p>0.05). Confirming the analysis above, and the report of Pan et al., post hoc comparisons (adjusted for age, time since injury, sex, and baseline CRS-R score) show that MCS- patients with evidence of CMD exhibit significantly greater gains than MCS- patients with no evidence of CMD (t(118)=3.062, p*_holm_*=0.019, d_mod_=1.259), with a trend towards significance also observed for VS patients with CMD compared to their counterparts with no evidence of CMD (t(118)=2.453, p*_holm_*=0.094, d_mod_=1.120), although MCS- patients with CMD did not appear to have significantly greater gains than VS patients with CMD (t(118)=0.882, p*_holm_*=0.76, d_mod_=0.508). Predictably, MCS+ patients exhibited greater CRS-R change at follow-up compared to both VS and MCS- patients with no evidence of CMD (t(118)=4.891, p*_holm_*<0.001, d_mod_=2.013 and t(118)=5.074, p*_holm_*<0.001, d_mod_=1.644, for VS and MCS- patients, respectively). Finally, and crucially, again no evidence was found that either group of patients with CMD had significantly different CRS-R gains compared to MCS+ patients (t(118)=1.620, p*_holm_*=0.539, d_mod_=0.893 and t(118)=0.839, p*_holm_*=0.760, d_mod_=0.385, for VS and MCS- patients, respectively; cf., Figure 1b).

Finally, repeating the analysis above on TBI patients only, thus restricting the sample to 24 no CMD, 17 CMD, and 53 MCS+ patients, returned the same pattern of results as in the main analysis. Specifically, the robust general linear model returned a significant effect of group (F(2,85)=18.269, p<0.001, ω^2^_p_=0.271) and initial CRS-R total score (F(1,85)=4.965, p=0.029, ω^2^_p_=0.059) on CRS-R change, with no significant effect of any other variable or interaction. Post-hoc pairwise comparisons again highlighted significant differences between the no CMD group and both the CMD (t(85)=3.765, p*_holm_*<0.001, d_mod_=1.705) and MCS+ (t(85)=5.309, p*_holm_*<0.001, d_mod_=1.778) groups, but no significant difference between the CMD and MCS+ groups (t(85)=0.143, p*_holm_*=0.887, d_mod_=0.074).

## Discussion

As discussed in Pan *et al.*^5^ and the connected commentary,^9^ the present analysis further confirms that the presence of CMD in patients presenting behaviorally as VS or MCS- is associated with better short-term outcomes compared to patients with the same behavioral phenotypes who show no evidence of CMD. Remarkably, however, and extending the science of DOC beyond what could be assessed by Pan and colleagues, the present work also shows that, at least in terms of short-term outcome (i.e., 3 months), patients with CMD are no different than MCS+ patients. As mentioned in the introduction, this finding is fairly intuitive. For, by virtue of showing a response to command, which implies the presence of language – along with all the many other ancillary cognitive processes needed to perform a complex mental task of the sort used in fMRI and EEG paradigms^27^ – patients with CMD are signaling that they are, in truth, MCS+ patients who were misdiagnosed as being VS or MCS-, likely due to the known limitations of behavior-based assessments.

This finding has a number of important implications. First, from the point of view of the nosology of DOC, the present findings suggest that, at least on the basis of short-term outcomes, VS and MCS- patients with CMD are best understood as misdiagnosed MCS+ patients, putatively due to the well known limitations intrinsic to behavioral assessments,^28^ and not a different category or diagnostic group. This conclusion strengthens the suggestion by the recent EU guidelines to diagnose DOC patients on the basis of their highest level of response across behavioral and non-behavioral (e.g., neuroimaging, electroencephalographic) assessments.^3^ By virtue of demonstrating the ability to perform response to command, patients incorrectly believed to be VS or MCS-, on the basis of behavioral responsiveness, are demonstrating they are, in fact, MCS+. While it might be extremely relevant, in the context of rehabilitation, to distinguish the mode by which a patient is able to demonstrate a state of MCS+ (e.g., by voluntarily moving their leg, hand, eyes, mouth, by engaging in a mental task, or by performing object recognition), the current data do not support this distinction being prognostically relevant and thus, at least in this respect, does not justify considering CMD as different clinical category compared to MCS+. In this sense, a patient appearing behaviorally VS or MCS- who is able to demonstrate response to command via advanced EEG-/MRI-based methods, might best be referred to as being “MCS+ with CMD,” to convey both their true level of consciousness and cognitive function as well as the absence of muscle-based voluntary responsiveness.

Second, the present findings also address the controversial issue of whether advanced imaging and electrophysiology assessments should be used in patients meeting behaviorally the criteria for MCS-,^21^ something which is endorsed by the European^3^ guidelines for management of patients with DOC, but not by the US^2^ ones. Given our data, the answer to this question is a clear yes, since patients who appear to be MCS- and have evidence of CMD show increases in CRS-R at follow-up greater than their counterparts with no evidence of CMD, and similar to those seen in MCS+ patients. This suggests that recent decisional instruments developed to guide ethical implementation of advanced techniques^21^ should be extended to also apply to MCS- patients. Finally, assessed at their broadest level, the present findings argue strongly against the view that more advanced brain imaging and electrophysiology techniques should not be “considered to be part of routine clinical practice,”^20^ as discussed previously.^19^

Finally, in evaluating the above results, a few important limitations need to be considered. First, the CMD patients were enrolled at a significantly later time post-injury, compared to the MCS+ patients (i.e., 3.51 months, compared to 1.58). This is a meaningful difference since time post-injury is known to be associated with early recovery in DOC patients,^29^ which could bias the present results. For this reason, all analyses include time since injury as a covariate and adjust for it in all inferential statistics, thus parceling out the effect of uneven time of enrollment. Second, another difference between the two samples included in the present analysis is the fact that while the time to follow-up in the patients described by Pan *et al.* was approximately 3 months, the average time between initial assessment and last follow-up in the patients described by Giacino, Whyte and collaborators was only 42.132 days (SD = 0.621). While this is a meaningful difference, it biases the data against the central thesis of this work – that CMD patients are in “just” MCS+ by another name. For, given the smaller interval between initial diagnosis and follow-up, MCS+ patients were effectively given less time to demonstrate changes in CRS-R, compared to the rest of the sample. Yet, as a group, they exhibited comparable gains to CMD patients. Third, the reader might know that the MCS+ sample was obtained from an interventional trial in some patients were administered Amantadine and the remainder were administered placebo. This might lead to the preoccupation that the CMD and MCS+ groups only appear to have similar short-term outcomes because a subset of MCS+ patients received a stimulant, thus enhancing their responsiveness. Yet, the central finding of Giacino, Whyte *et al*. is that Amantadine hastens recovery, but does not lead to group differences (i.e., amantadine versus placebo) when comparing, as done here, the first and last assessments.^11^ The administration of Amantadine in a subset of the patients thus cannot be seen as leading to the observed results. Last, another notable confounding variable in the present analysis is the fact all 53 MCS+ patients suffered a traumatic brain injury (TBI), as per the inclusion criteria of Giacino, Whyte and colleagues,^11^ whereas the CMD and no CMD patients included a mix of traumatic (52%), cardiovascular (26%), and anoxic (22%) brain injuries. While traumatic injuries are known to be associated with better outcomes,^18^ restricting the analysis of CRS-R total score change to TBI patients only (across all three groups; i.e., no CMD, CMD, MCS+) leads to the same pattern of results, suggesting that this imbalance in sample etiology is not driving the reported findings.

In conclusion, the present work suggests that, at least in terms of short-term outcomes, CMD patients are, in terms of short-term outcomes, MCS+ patients by another name. In addition, the present results further stress the importance of advanced imaging and electrophysiology assessments in appropriately diagnosing DOC patients, including not only patients appearing entirely behaviorally unresponsive (i.e., VS), but also patients meeting behaviorally criteria for MCS-. Re-assessment of the question addressed here with longer-term outcomes will be important to confirm and extend the current conclusion.

## Acknowledgements

The author would like to acknowledge Dr. Giacino, Dr. Whyte, and collaborators for providing longitudinal data for MCS+ patients collected as part of their prior published work.

## Supplementary material

Supplementary material is available at *Brain* online.

