## Appendix for "Is cognitive motor dissociation just a minimally conscious state “plus” by another name?"

Martin M. Monti [1,2]

**Author affiliations:**

1 Department of Psychology, University of California Los Angeles, Los Angeles, CA 90095, USA

2 Brain Injury Research Center, Department of Neurosurgery, University of California Los Angeles, Los Angeles, CA 90095, USA

Correspondence to: Martin M. Monti

Full address: UCLA Department of Psychology, Los Angeles, CA 90095, USA

**Running title**: Is CMD just MCS+ by another name?

**Keywords:** separate by semi-colon; 12 point; maximum of six; do not use words already mentioned in the title or abstract

### Appendix

**Table A1**. Demographics and clinical information for the of the sample for each of the 5 groups included in the more granular analysis. (Abbreviations are the same as in Table 1.)

| | Group | N | Mean | Med | SD | Min | Max |
|---|---|---|---|---|---|---|---|
| **Age (years)** | **VS no CMD** | 27 | 39.074 | 37.00 | 14.848 | 15.000 | 66.00 |
| | **VS with CMD** | 18 | 39.056 | 41.50 | 13.523 | 19.000 | 62.00 |
| | **MCS- no CMD** | 17 | 35.824 | 37.00 | 15.864 | 16.000 | 70.00 |
| | **MCS- with CMD** | 16 | 36.688 | 41.00 | 13.375 | 15.000 | 52.00 |
| | **MCS+** | 53 | 38.338 | 38.03 | 16.409 | 16.263 | 64.80 |
| **TSI (mo)** | **VS no CMD** | 27 | 4.481 | 3.50 | 3.781 | 1.000 | 20.00 |
| | **VS with CMD** | 18 | 3.194 | 3.00 | 1.446 | 1.000 | 6.00 |
| | **MCS- no CMD** | 17 | 3.735 | 3.00 | 2.857 | 1.500 | 14.00 |
| | **MCS- with CMD** | 16 | 3.875 | 2.25 | 4.031 | 1.500 | 18.00 |
| | **MCS+** | 53 | 1.577 | 1.33 | 0.673 | 0.900 | 3.57 |
| **CRS-R total (baseline)** | **VS no CMD** | 27 | 5.778 | 6.00 | 1.281 | 4 | 9 |
| | **VS with CMD** | 18 | 6.056 | 6.50 | 1.162 | 3 | 7 |
| | **MCS- no CMD** | 17 | 8.941 | 8.00 | 1.638 | 7 | 12 |
| | **MCS- with CMD** | 16 | 9.375 | 9.00 | 1.708 | 7 | 13 |
| | **MCS+** | 53 | 13.170 | 13.00 | 3.683 | 6 | 20 |
| **CRS-R total (follow-up)** | **VS no CMD** | 27 | 6.667 | 7.00 | 1.754 | 4 | 10 |
| | **VS with CMD** | 18 | 11.500 | 10.50 | 4.423 | 6 | 23 |
| | **MCS- no CMD** | 17 | 10.118 | 9.00 | 2.913 | 7 | 17 |
| | **MCS- with CMD** | 16 | 16.063 | 18.00 | 4.553 | 7 | 22 |
| | **MCS+** | 53 | 19.302 | 22.00 | 5.301 | 2 | 23 |
| **CRS-R change** | **VS no CMD** | 27 | 0.889 | 0.00 | 1.553 | 0 | 6 |
| | **VS with CMD** | 18 | 5.444 | 4.00 | 4.706 | 1 | 20 |
| | **MCS- no CMD** | 17 | 1.176 | 0.00 | 2.721 | 0 | 10 |
| | **MCS- with CMD** | 16 | 6.688 | 9.00 | 4.207 | 0 | 12 |
| | **MCS+** | 53 | 6.132 | 6.00 | 5.299 | -14 | 15 |
